# Shared genetics between breast cancer and its predisposing diseases identify novel breast cancer treatment candidates

**DOI:** 10.1101/2024.05.14.24307374

**Authors:** Panagiotis N. Lalagkas, Rachel D. Melamed

## Abstract

Breast cancer is a major public health issue. Current treatment options, while effective, have severe side effects. FDA-approved drugs have known safety and pharmacological profiles and some, such as metformin, have been tested in clinical trials for repurposing for breast cancer. However, clinical trials are slow and expensive, which creates the need for innovative approaches to accelerate drug repurposing for breast cancer. We have previously shown that genes associated with Mendelian diseases that predispose patients to certain cancers are enriched for successful drug targets, due to their pleiotropic effects. Here, we extend our approach to exploit clinical associations between breast cancer and its predisposing diseases for drug repurposing. We hypothesize that pleiotropic genes shared between breast cancer and its predisposing diseases can help us discover new uses for drugs currently approved only for the predisposing diseases. To test our hypothesis, we compile a list of six traits known to increase breast cancer risk (predisposing diseases). Using GWAS summary statistics and local genetic correlation analysis, we find 84 genomic loci harboring mutations with positively correlated effects between breast cancer and each predisposing disease. These loci contain 194 protein-coding genes (shared genes). Using a network biology approach and canonical pathways, for each disease pair, we connect drugs already indicated for the predisposing disease to its shared biology with breast cancer and identify drug repurposing candidates for breast cancer. Finally, we show that our list of candidate drugs is enriched for currently investigated and indicated drugs for breast cancer (OR=9.28, p=7.99e-03). Our findings suggest a novel way to accelerate drug repurposing for complex diseases by leveraging shared genetics.

## Introduction

Breast cancer is a major public health issue and the most common type of cancer in women worldwide^1^. Current treatment options, such as chemotherapy, hormone therapy, and immunotherapy^2^, while effective, often have severe side-effects that impair patients’ quality of life and adherence to treatment^3,4^. On the other hand, existing FDA-approved drugs have established safety and pharmacological profiles, and testing them for repurposing for breast cancer treatment has been an attractive strategy^5^. For instance, metformin, the most commonly prescribed anti-diabetic drug, has shown anti-proliferative effects in pre-clinical studies and is currently undergoing clinical trials for breast cancer treatment. This surprising effect may be explained by its downstream effects activating *AMPK* protein kinase, a tumor suppressor critical in regulating cell proliferation^6,7^. However, oncology clinical trials are slow, expensive and have up to 95% attrition rates, mainly due to lack of understanding of the mechanism of action of the tested drug prior to the clinical trial^8–10^. Therefore, new approaches are needed to both accelerate drug repurposing for breast cancer treatment and provide biological insights to design clinical trials with higher success rates.

Genome-wide association studies (GWAS) associate genes with a disease; drugs targeting those genes are twice as likely to successfully treat that disease^11–13^. For example, a recent breast cancer GWAS found a strong signal in the *ESR1* locus which is the target of tamoxifen, a widely used drug for breast cancer treatment^14^. This highlights the great potential of genetics in informing drug discovery and repurposing^15^. However, the inherent limitations of GWAS, such as the need for large sample sizes and the challenge of identifying true causal genes, limit their ability to inform drug repurposing^16,17^. This creates the need for innovative ways to build on genetics findings to discover drugs for diseases of high public importance, such as breast cancer.

To this end, we propose that *shared genetics between breast cancer and other diseases* can inform drug repurposing for breast cancer. This idea is supported by prior research. For one, it has been shown that clinically co-occurring diseases share genetics^18,19^. This suggests that shared biological processes may explain why a health condition might predispose individuals to breast cancer. Consequently, drugs approved for such predisposing diseases and targeting shared biology with breast cancer might hold potential for breast cancer treatment. In further support of this concept, we have previously built on work showing bearers of Mendelian disease mutations may suffer increased risk of certain cancers, due to a hidden role of the Mendelian disease genes in the comorbid cancer. In a follow-up study, we showed that the Mendelian disease genes implicated for a cancer are enriched for successful drug targets^20^. Building on these observations, here, we propose the repurposing of drugs based on shared genetic etiology.

Ultimately, we propose 59 drugs that have not been previously tested for breast cancer, accompanied by biological insights supporting their potential therapeutic effect on breast cancer.

## Results

### Overview of approach

Health conditions including high cholesterol and type 2 diabetes incur increased risk for breast cancer. Previous genetics research has supported a biological explanation: these diseases, which we will refer to as *predisposing diseases*, share genetic variation with breast cancer. Building on this, we hypothesize that finding the shared genetics between breast cancer and its predisposing diseases can help us discover new drugs for breast cancer. Specifically, we hypothesize that drugs approved for a predisposing disease and targeting its shared biology with breast cancer can treat the latter disease (**Figure 1A**).

**Figure 1.**
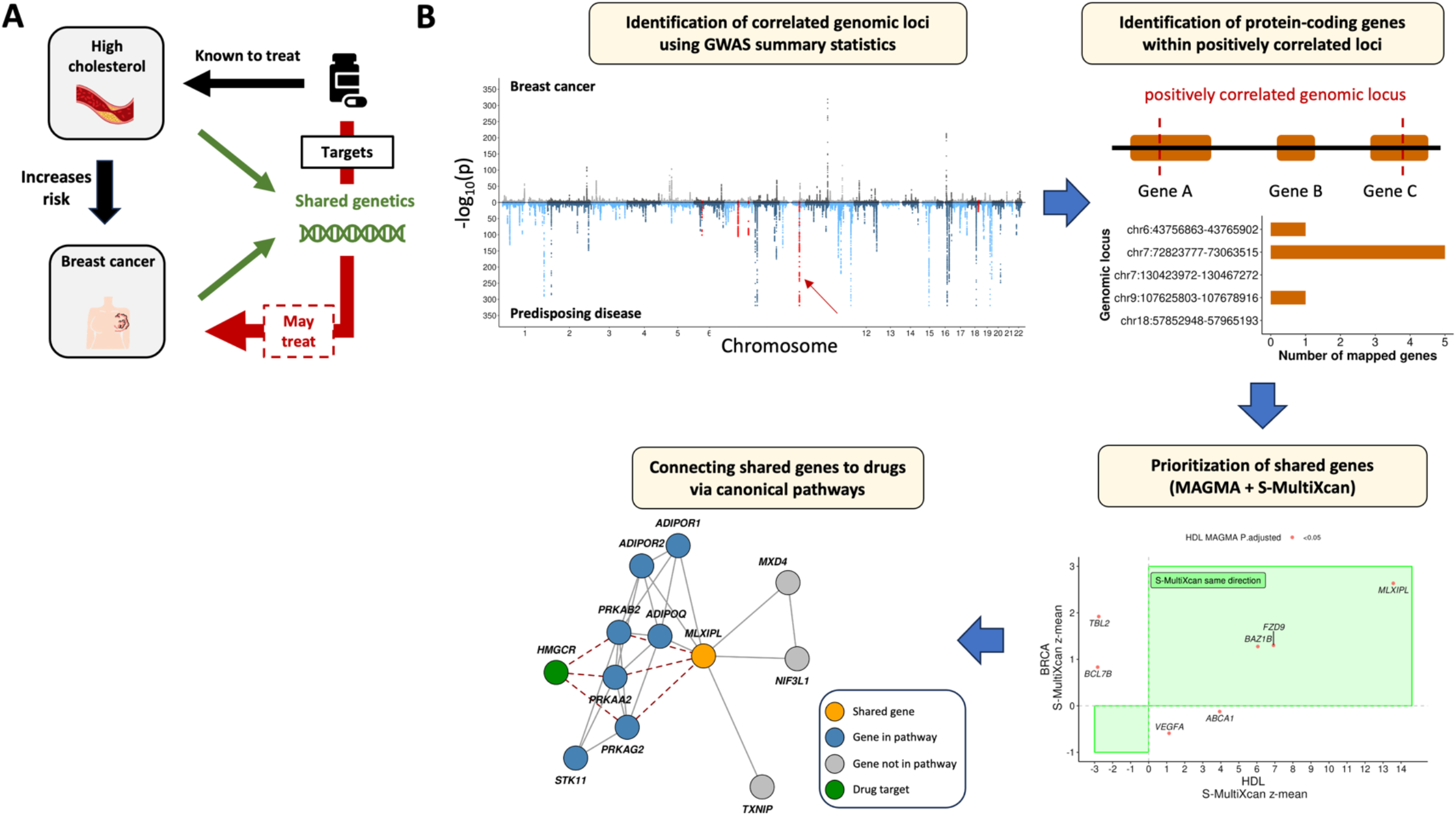
Outline of the approach. **A**. Tested hypothesis that a drug treating a health condition known to increase the risk for breast cancer and targets the shared biology of both conditions may also treat breast cancer. **B**. Proposed workflow to identify the most likely shared genes between breast cancer and a predisposing disease and connect them to drugs approved for the predisposing disease.

To test this hypothesis, we first search the scientific literature (epidemiological and statistical genetic studies) for diseases with genetic variation known to predispose individuals to breast cancer. We find six such diseases (**Table 1**).

**Table 1.**
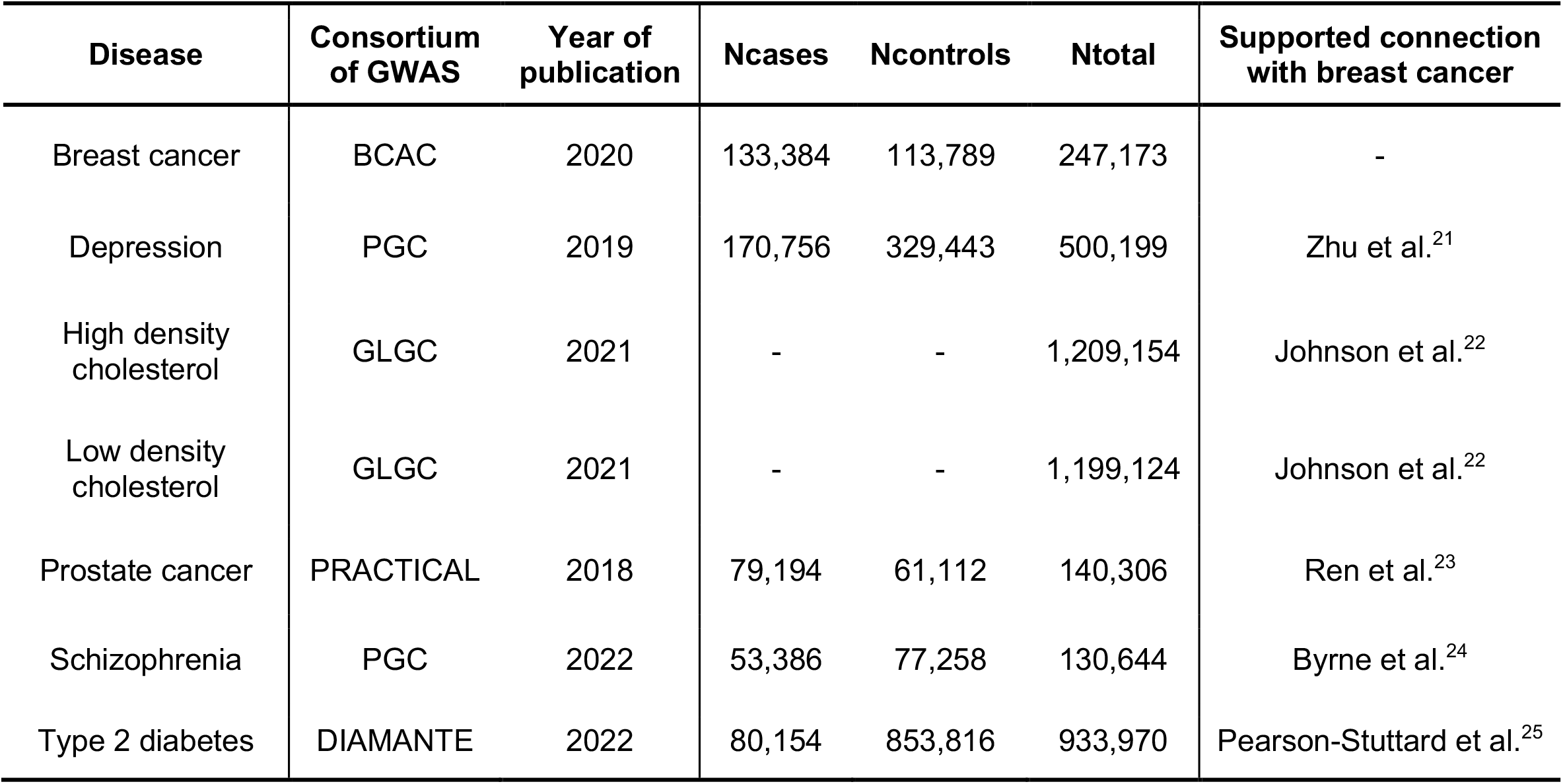
Evidence for shared etiology of predisposing conditions, and genome-wide association study (GWAS) summary statistics for breast cancer and its six predisposing diseases used in this study. All GWAS include samples of European ancestry. BCAC: Breast Cancer Association Consortium; PGC: Psychiatric Genomics Consortium; UKBB: United Kingdom BioBank; GLGC: Global Lipids Genetics Consortium; PRACTICAL: Prostate Cancer Association Group to Investigate Cancer-Associated Alterations in the Genome; DIAMANTE: Diabetes Meta-Analysis of Trans-Ethnic association studies

Next, we aim to identify the shared genetics between each pair of breast cancer and a predisposing disease that can be tied to medications currently used to treat the predisposing disease. We use publicly available GWAS summary statistics data (**Table 1**) and a local genetic correlation analysis, to find the shared genetics. Following the identification of likely shared genes for each disease pair, we connect them to drugs currently used to treat the predisposing disease. Because we wish to both make this connection and gain interpretation about the pathways involved, we link each shared gene, and each set of drug targets, to canonical pathways using a network biology approach. By finding drugs that target shared pathways, we both prioritize candidate drugs for repurposing for breast cancer and provide biological insights that support their effect in disease treatment. **Figure 1B** illustrates the complete workflow using the example of high HDL and breast cancer.

Finally, we evaluate our list of candidate drugs. To do so, we compile a list of 583 drugs either in clinical trials (N=451) or approved (N=132) for breast cancer and test for enrichment within our candidate drugs.

### Discovering shared genetics between breast cancer and its predisposing diseases

The first step in testing our hypothesis is to identify genomic loci likely to drive the shared etiology between breast cancer and a predisposing disease. Although cross-trait Linkage Disequilibrium Score Regression (LDSC) is a widely adopted method to identify genetic correlation between a pair of phenotypes using GWAS summary statistics, its genome wide nature means it does not provide insight into particular genes driving the correlation. To gain that insight, we perform a local genetic correlation analysis for each pair of breast cancer and predisposing disease, using LOGODetect^26^. Across all disease pairs, we identify 59 negatively (per disease pair: median=5.5; min=0; max=34) and 84 positively (per disease pair: median=10; min=5; max=37) correlated genomic loci (**Figure 2A, Supplementary Tables 1 & 2**). Notably, the identified loci are distributed across all autosomal chromosomes and not localized in specific parts of the genome (**Figure 2B**). In order to confirm that LOGODetect does not discover shared loci for disease pairs without known epidemiological or clinical associations, such as high LDL-depression and high LDL-schizophrenia, we repeat the analysis for these pairs. LOGODetect does not identify any significant correlated loci.

**Figure 2.**
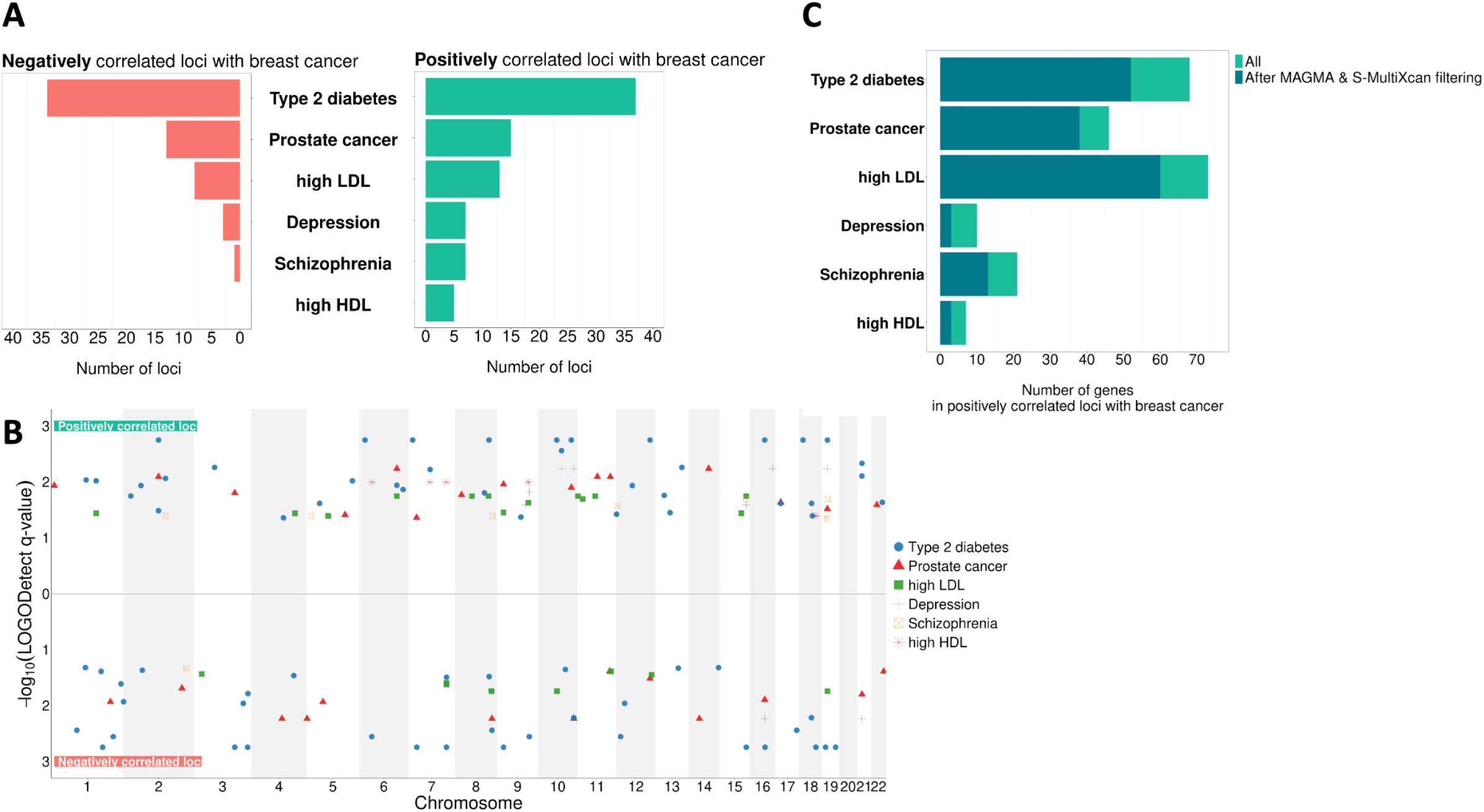
Shared genetics between breast cancer and its predisposing diseases. **A**. Number of negatively and positively correlated loci between each breast cancer and predisposing disease pair. **B**. Position in the genome of the identified correlated loci for each breast cancer and predisposing disease pair. The y-axis represents the -log10(q-value) of each locus as provided by LOGODetect. **C**. Number of protein-coding genes located within the positively correlated loci for each breast cancer and predisposing disease pair, before and after applying the MAGMA & S-MultiXcan filters.

Next, for each predisposing disease, we seek to prioritize genes in the correlated genomic loci that are the most likely drivers of shared etiology with breast cancer. Since our ultimate goal is to recommend candidate drugs for repurposing for breast cancer, we are more interested in genes exhibiting effects in the same direction in both breast cancer and predisposing disease. Therefore, for the downstream analysis, we focus on genomic loci found to be positively correlated for each disease pair. Using the SNP2GENE function from FUMA^27^, we extract a total of 194 protein-coding genes that fall within all the positively correlated loci (median of 37 protein-coding genes per disease pair; min=8; max=81). For each disease pair, these genes constitute the list of shared genes that likely drive the shared etiology with breast cancer.

From this list, we seek the subset most likely to contribute to druggable shared etiology. That is, we wish to prioritize genes more likely to 1) participate in shared pathophysiological processes and 2) relate to the effect of drugs on the predisposing disease. First, we use MAGMA, a tool that aggregates the effect of all SNPs within a gene, to keep genes significantly associated with the predisposing disease in every disease pair^28^. This filter is important as the predisposing disease increases the risk for breast cancer, implying that any genes underlying that shared risk should, at a minimum, impact the predisposing disease. Second, we use S-MultiXcan, a tool that uses GWAS summary statistics data to genetically predict the expression of a gene summarized across 49 GTEx tissues^29^. We keep only genes with genetically predicted expression consistently aligned in the same direction (either downregulated or upregulated) in both diseases in a disease pair, as dysregulation in opposite directions is not easily interpretable and not useful for therapeutic purposes. For instance, *ABCA1* is found to be one of the genes shared between high LDL and breast cancer. Probucol, an approved anti-cholesterol drug, inhibits *ABCA1*. But, *ABCA1* is upregulated in high LDL and downregulated in breast cancer, suggesting that inhibitions of this gene might not lead to desired treatment outcome for breast cancer^30^. **Figure 2C** shows the number of shared genes within positively correlated loci between breast cancer and each predisposing disease, before and after applying the filters. The full list of shared genes for each disease pair is provided in **Supplementary Table 3**.

### Connecting shared genetics to drugs

The next step is to connect the identified shared genes to drugs treating the predisposing diseases. We make this connection through canonical pathways. In that way, we can capture the biological processes which 1) are involved with identified shared genes and 2) are related to drugs indicated for the predisposing disease. First, for each breast cancer and predisposing disease pair, we connect the identified shared genes to canonical pathways using the STRING protein-protein interaction network and a network propagation algorithm (**Supplementary Table 4**). Notably, our workflow identifies pathways with known roles in tumorigenesis and progression that are also disrupted in the breast cancer predisposing diseases (**Figure S1**). For instance, we identify the *WNT signaling pathway* to be part of the shared etiology between breast cancer^31^and each of high HDL^32^, prostate cancer^33^and type 2 diabetes^34^. Additionally, the *mTOR* signaling pathway, another cancer-related pathway^35,36^, is found to be shared between breast cancer and high LDL^37^.

After connecting the shared genes for every disease pair to pathological processes, we seek to find which drugs target them. To do so, we use a similar approach and connect targets of a drug currently approved for a predisposing disease to the shared canonical pathways between breast cancer and that disease (**Figure 3, Supplementary Table 5**). Drugs significantly connected to at least one shared canonical pathway are considered candidate drugs for repurposing for breast cancer.

**Figure 3.**
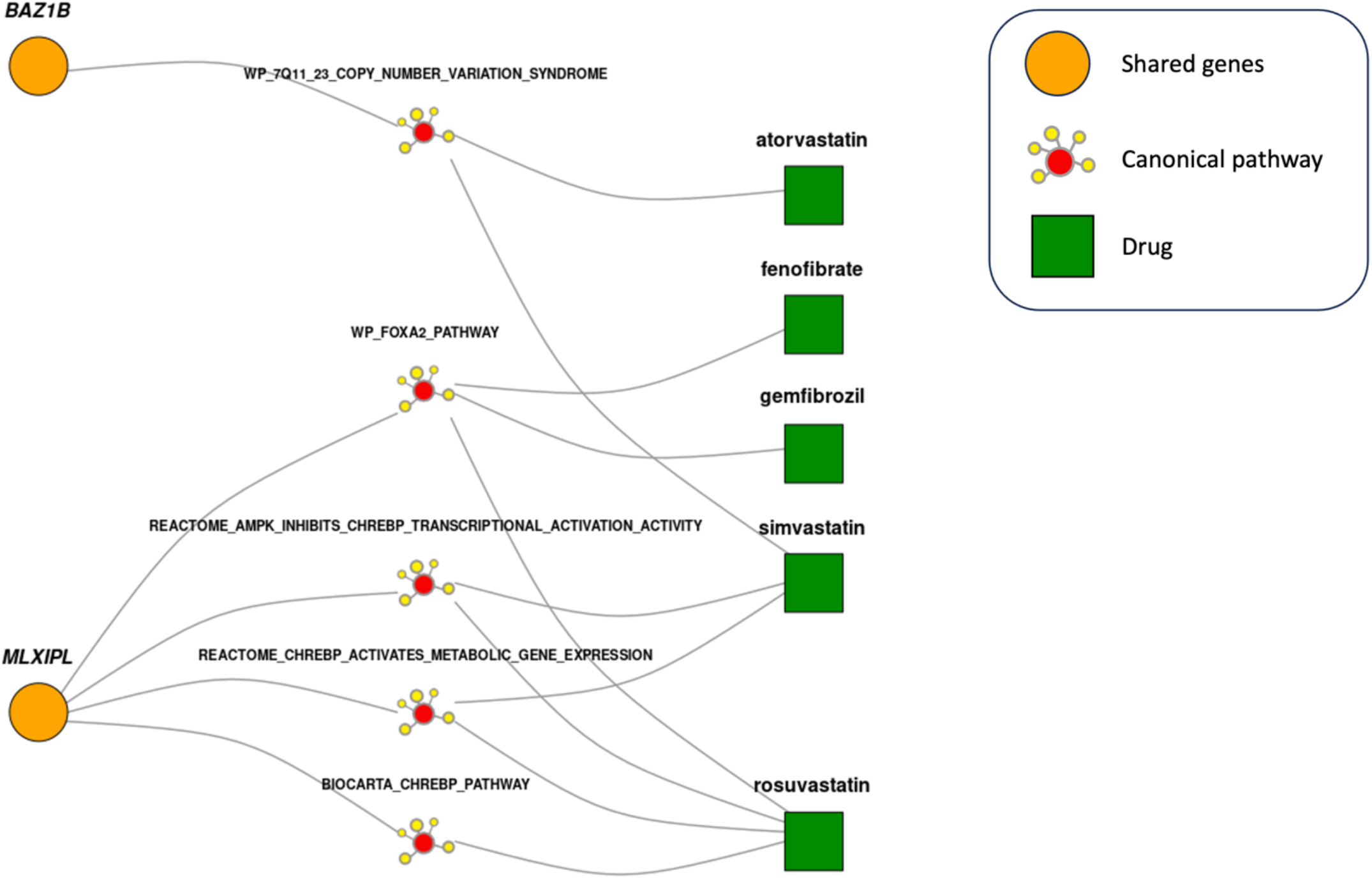
Drugs treating high HDL and targeting its shared biology with breast cancer. These drugs are recommended for repurposing for breast cancer and the shared pathways they target provide a biological basis for the repurposing. Only canonical pathways significantly linked to both identified shared genes and high HDL drugs are shown.

### Evaluation of drug repurposing and prioritization of new candidates

The final step is to assess the efficacy of our approach in identifying promising candidate drugs for repurposing for breast cancer. To do so, we compare our list of candidate drugs to those currently under investigation or approved for breast cancer treatment. In total, out of 112 approved drugs for the six breast cancer predisposing diseases, 16 have undergone testing or received approval for breast cancer treatment (**Figure 4A**). Remarkably, our method identifies 15 out of these 16 drugs, while also offering insights into the specific genes and pathways that might explain their effect on breast cancer (**Supplementary Table 5**). When considering all the recommended candidate drugs for repurposing, we find a significant enrichment for drugs currently investigated or approved for breast cancer (OR=9.28, p=7.99e-03, one-sided Fisher’s exact test).

**Figure 4.**
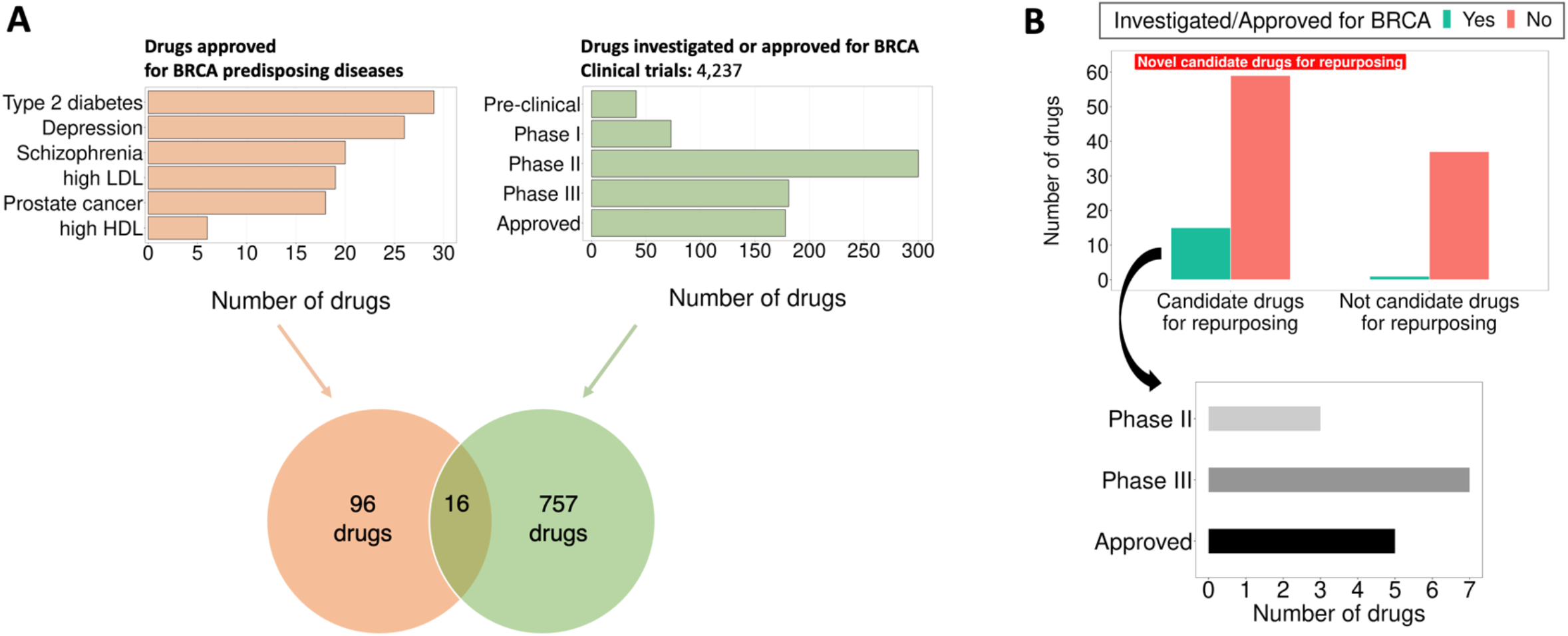
Evaluation of recommended candidate drugs for repurposing in breast cancer. **A**. Out of all drugs approved for breast cancer predisposing diseases, sixteen have been investigated or approved for breast cancer treatment. **B**. Our recommended candidate drugs for repurposing in breast cancer treatment are significantly enriched for drugs already investigated or approved for breast cancer (OR=9.28, p=7.99e-03, one-sided Fisher’s exact test). Notably, our list of candidate drugs includes 15 of the 16 drugs currently in advanced clinical trials (Phase II or III) or approved for breast cancer treatment. BRCA: breast cancer

Our workflow identifies *HMGCR* to be shared between high LDL and breast cancer. However, *HMGCR* is also the target of statins, a group of drugs approved for lowering the LDL blood levels. Therefore, to ensure that the significance of our results is not driven by this instance where a shared gene is also the target of an approved drug for the predisposing disease, we repeat the analysis by excluding the high LDL-breast cancer pair. Again, we find strong enrichment (OR=9.35, p=9.66e-03, one-sided Fisher’s exact test), highlighting the ability of our approach in connecting shared biology to drugs even when a shared gene is not directly targeted by a drug.

In conclusion, we recommend 76 candidate drugs for repurposing while providing biological insights (specific genes and pathways) supporting their potential use in breast cancer treatment. Among these candidates, 15 are already in advanced clinical trial phases or approved for breast cancer treatment, while 59 are novel candidate drugs (**Figure 4B**).

## Discussion

In our previous work, we exploited monogenic diseases that predispose their bearers to complex diseases, showing that drugs targeting the causal gene of the monogenic disease are good candidates for treating the associated complex disease^20^. With that study we introduced the idea that predisposing health conditions of a complex disease, and their genetics, can be used to inform drug repurposing for the complex disease. Building on that work, here, we develop an approach to investigate whether complex disease risk factors that predispose individuals to a complex disease can inform the repurposing of existing drugs for the complex disease. To test this, we use breast cancer as an example of a well-studied and common complex disease, with many predisposing risk factors. We show that shared genes and pathways between breast cancer and its predisposing diseases can point to promising candidate drugs for repurposing for breast cancer treatment that target the shared pathways. Importantly, our recommended candidate drugs are enriched for those currently undergoing clinical trials or already approved for breast cancer treatment, highlighting the ability of our approach in identifying likely successful candidate drugs for repurposing.

Pleiotropy has been previously used for the identification of shared biology between diseases from the same body systems, such as psychiatric diseases. However, our study aims to identify shared genetic factors between breast cancer and its predisposing diseases which primarily affect different body systems, such as depression and high HDL. Despite affecting distinct anatomical regions, these disease pairs share pathophysiological processes which suggests that association is not just due to the known similarity between the phenotypes. Our approach identifies these pleiotropic genes using a local genetic correlation analysis and links them to canonical pathways which are used to prioritize candidate drugs for repurposing. With that, our study is the first to propose an alternative way to analyze publicly available GWAS summary data that can both suggest new uses for existing drugs and point towards the genes and pathways supporting drug repurposing.

We present an example to illustrate the power of our approach in both identifying shared biology between diseases and providing a biological basis for the repurposing of a drug. It is known that elevated levels of HDL increase the risk for breast cancer^22^. By analyzing this pair of diseases, we identify *MLXIPL* as a shared gene between breast cancer and high HDL. We also find that *MLXIPL* is significantly connected to the *FOXA2* pathway. Interestingly, the *FOXA2* pathway is known to play a role in both breast cancer pathogenesis and lipid metabolism (pleiotropic effect)^38–40^. Among the approved lipid-lowering medications, we find two fibric acids (fenofibrate and gemfibrozil) and one statin (rosuvastatin) significantly connected to the *FOXA2* pathway. Notably, both drug categories have demonstrated anti-cancer properties in preclinical and clinical studies, respectively^41,42^. This example shows that our approach can identify meaningful biological signals. It also shows that by leveraging the shared biology between breast cancer and its predisposing factors, we can prioritize candidate drugs for repurposing while also providing plausible biological mechanisms through which these drugs may impact breast cancer.

Our approach has some limitations. First, we define shared genes as those located within the detected, positively correlated shared genomic loci for each disease pair. However, a SNP in a shared locus might have distal regulatory effects on genes located outside that locus and including those genes could potentially result in a more complete list of shared genes for each disease pair. Second, we rely on pleiotropy to identify shared genetic factors between breast cancer and its predisposing diseases. However, it is possible that a disease might increase the risk for breast cancer through indirect or interaction effects, which are not captured by our approach. Third, redundant canonical pathways in the Molecular Signatures Database (MSigDB) could result in the identification of fewer significantly shared pathways between diseases than their actual number due to multiple testing corrections. Fourth, we analyzed GWAS summary statistics data only from the European population, due to greater sample sizes and data availability. Future studies analyzing data from diverse populations may discover shared loci missing from our analysis.

In conclusion, our approach detects and leverages the shared biology between pairs of breast cancer with its predisposing diseases to suggest novel drugs for breast cancer treatment, while also providing a biological basis for each drug recommendation. Future work can exploit our list of candidate drugs for repurposing and evaluate their efficacy in experimental settings. While we specifically applied our approach to the case of breast cancer, we trust that it can be applied to recommend novel candidate drugs for repurposing for any complex disease with a known set of predisposing diseases and available GWAS summary statistics. Therefore, it serves as a valuable tool for not only identifying promising candidate drugs for repurposing across various complex diseases, but also suggesting the disease networks involved in repurposing.

## Methods

### Data collection and preprocessing

#### Genetics data

We download the most recent, publicly available GWAS summary statistics data of European ancestry for breast cancer^43^, depression^44^, high HDL^45^, high LDL^45^, prostate cancer^46^, schizophrenia^47^ and type 2 diabetes^48^. For each GWAS, we keep only bi-allelic SNPs in autosomal chromosomes that match to unique rsIDs. We also harmonize the effect and alternate allele for each SNP using the 1K Genomes reference panel of Europeans. The GWAS for schizophrenia included an imputation quality score for each SNP and we filter for SNPs with imputation score ≥0.3.

#### Protein-protein interaction network

We download the STRING protein-protein interaction (PPI) network for humans (version 11.5, date of download: May 8, 2023). This network contains both physical and functional interactions of proteins and assigns a confidence score to each interaction (edge) that represents the amount of evidence supporting it. We use this score to filter for edges with high confidence (score ≥ 0.7). We also remove multiple edges and loops and map each gene-node to entrezIDs using a file provided by STRING (“9606.protein.aliases.v11.5.txt”). The final PPI network contains 16,115 nodes and 240,541 edges.

#### Canonical pathways

We download a list of gene sets for canonical pathways from the MSigDB (version 2023.2). This is a list of manually curated gene sets by domain experts and includes information from five databases: BioCarta, KEGG (Kyoto Encyclopedia of Genes and Genomes), Reactome, PID (Pathway Interaction Database) and WikiPathways. For each canonical pathway, we filter for genes found in the STRING PPI network. Eventually, we have a list of 3,795 canonical pathways that we use to connect shared genes to drugs.

#### Drugs indicated for breast cancer predisposing diseases

For each of the six breast cancer predisposing diseases (depression, high HDL, high LDL, prostate cancer, schizophrenia, type 2 diabetes), we compile a list of FDA-approved drugs using RxNORM and annotate them with gene-targets from DrugBank (version 5.19, date of download: June 16, 2022). We do not filter the list of targets based on pharmacological action. Finally, we keep the drug gene-target with unique entrezIDs that are found in the STRING PPI network.

#### Drugs investigated or indicated for breast cancer

To find drugs currently investigated for breast cancer, we download clinical trial data from the Aggregate Content of ClinicalTrials.gov (AACT) database in a pipe-delimited format (date of download: November 4, 2022; note that it is updated daily). AACT is a publicly available relational database that contains information about all the studies registered in ClinicalTrials.gov. We obtain information for 432,597 clinical trials that were registered in ClinicalTrials.gov by the date of download. Using a manually curated list of MeSH codes related to breast cancer (D001943, D000071960, D018270, D018275, D061325, D058922, D064726, D000069584), we keep 4,237 clinical trials that tested 774 drugs for breast cancer treatment.

To find drugs currently indicated for breast cancer, we obtain the indications of all FDA-approved drugs from RxNORM and filter for those that treat breast cancer using the MeSH terms mentioned above. We find 68 breast cancer indicated drugs and combine them with the investigated drugs to get a complete list of drugs for breast cancer. Again, all drugs are coded in DrugBank IDs.

### Finding likely shared genes between breast cancer and its predisposing diseases

To identify shared genes between each pair of breast cancer and predisposing diseases, we use GWAS summary statistics data and LOGODetect (default settings) to perform a local genetic correlation analysis. LOGODetect scans the entire genome to identify loci with correlated SNP effects in two GWAS, accounting for linkage disequilibrium. Following the identification of shared loci for each disease pair, we use the FUMA SNP2GENE function to compile a list of protein-coding genes positionally located within and ±10 kb upstream and downstream of positively correlated loci (default setting). We do this by providing to FUMA SNP2GENE a list of all SNPs within the identified loci and not only the genome-wide significant ones. Eventually, for each breast cancer and predisposing disease pair, we obtain a list of protein-coding genes (entrezIDs) that are part of their shared etiology (termed as shared genes).

We then seek to prioritize genes that are more likely to contribute to the shared biology for each disease pair. To achieve this, we utilize two commonly used tools: MAGMA (gene-based) and S-MultiXcan (cis-eQTL based). First, we run MAGMA with default settings to obtain gene-based p-values for each disease and adjust them for multiple hypothesis testing using the Benjamini-Hochberg correction method. Second, we use S-MultiXcan to obtain the genetically predicted expression of genes, summarized across 43 GTEx tissues. Then, for each disease pair, we filter the previously identified shared genes for those that are significantly associated with the predisposing disease (MAGMA p.adjusted<0.05) and are predicted to be dysregulated in the same direction in both diseases (sign of S-MultiXcan calculated z-score). Ultimately, these two filtering steps, one gene-based and one cis-eQTL-based, help us prioritize shared genes more likely to contribute to the shared biology of a breast cancer and predisposing disease pair.

### Connecting shared genes to drugs via canonical pathways

Our goal here is to link the shared genes with drugs and gain insights into the implicated biological pathways. To do so, we first find which canonical pathways are significantly connected to the shared genes using the STRING PPI network and the Personalized PageRank (PPR) algorithm. PPR uses a user-defined gene, named seed gene, as a starting point and performs random walks in the PPI network, with a probability (default=0.8) of returning to the seed gene after each step. This process assigns a score to each gene based on its connectivity with the seed gene: genes with higher connectivity receive higher scores. Using each shared gene for a disease pair as a seed gene, we calculate the average PPR score of genes within a canonical pathway. To determine if the observed score is higher than expected by chance, we compare it to random PPR scores obtained through 1,000 permutations (more details in “Statistical analysis - Permutation tests”). Ultimately, for each breast cancer and predisposing disease pair, this process yields a list of canonical pathways that are significantly connected (p_permutation_<0.05) to the previously identified shared genes (referred to as shared canonical pathways).

We then seek to link the shared canonical pathways to drugs approved for a predisposing disease. To do so, we repeat the above analysis but this time, we use a target of a drug as a seed gene (instead of a shared gene). Similarly, we calculate the average PPR score of genes within a shared canonical pathway and compare it to average PPR scores obtained after 1,000 permutations. Drugs approved for a predisposing disease and significantly connected to a shared canonical pathway (p_permutation_<0.05), are considered to target the shared etiology with breast cancer. Therefore, these drugs constitute the prioritized candidate drugs for repurposing for breast cancer, and the shared genes and canonical pathways they target provide biological insights for their effect on breast cancer.

### Statistical analysis

#### Evaluating candidate drugs for repurposing

We find 76 unique drugs approved for a predisposing disease that can be linked with its shared biology with breast cancer. Using these drugs alongside those currently in clinical trials or approved for breast cancer treatment, we test for significant overlap using the Fisher’s one-sided exact test.

#### Permutation tests

To assess the significance of the observed associations between shared genes, drugs, and canonical pathways, we conduct permutations tests.

First, for each breast cancer and predisposing disease pair, we connect each canonical pathway to the identified shared genes. To do so, we use each shared gene as a seed gene, and we calculate the average PPR score for the genes in a canonical pathway. To determine the significance of these connections, we create a null distribution of average PPR scores for genes within a canonical pathway. This involves grouping all genes in the PPI network into four bins based on their degree (quantiles). Subsequently, for each seed (shared) gene, we randomly select 1,000 genes from the same degree bin and calculate the average PPR score for genes within the canonical pathway. These scores are then compared to the observed average PPR score, and a permuted p-value is calculated based on how often the observed score is lower than the permuted scores. This process yields a permuted p-value for every canonical pathway-shared gene pair, which is then adjusted for the number of shared genes and canonical pathways tested using the Bonferroni correction method.

Second, we link the shared canonical pathways to drugs approved for a predisposing disease to prioritize candidate drugs for repurposing. To do so, we follow a similar approach as above, but this time, we use random genes matched to the gene-target of each drug using the four degree bins. This process yields a permuted p-value for every shared canonical pathway-drug target pair, which is then adjusted for the number of drug targets tested using the Bonferroni correction method.

## Data Availability

The data are all publicly available at GWAS catalog.

