## Supplementary Figure for "Shared genetics between breast cancer and its predisposing diseases identify novel breast cancer treatment candidates"

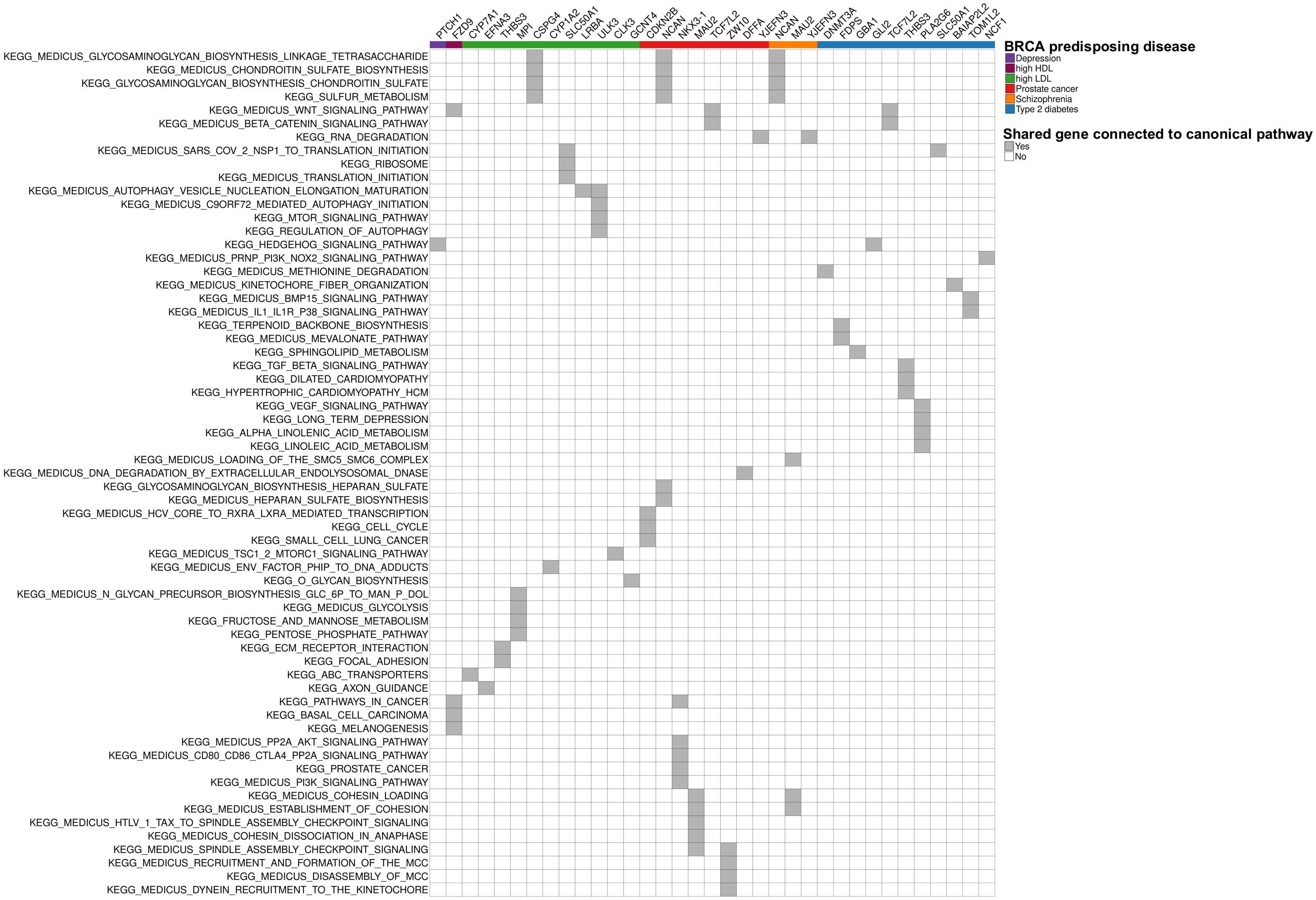


**Supplementary Figure 1.** KEGG canonical pathways connected to the shared genes between breast cancer and its predisposing diseases.
